# Expansion of Artemisinin Partial Resistance Mutations and Lack of Histidine Rich Protein-2 and -3 Deletions in *Plasmodium falciparum infections* from Rukara, Rwanda

**DOI:** 10.1101/2023.12.17.23300081

**Authors:** Cecile Schreidah, David Giesbrecht, Pierre Gashema, Neeva Young, Tharcisse Munyaneza, Claude Mambo Muvunyi, Kyaw Thwai, Jean-Baptiste Mazarati, Jeffrey Bailey, Jonathan J Juliano, Corine Karema

## Abstract

**Background:** Emerging artemisinin resistance and diagnostic resistance are a threat to malaria control in Africa. *Plasmodium falciparum* kelch13 (K13) propeller-domain mutations that confer artemisinin partial resistance have emerged in Africa. K13-561H was initially described at a frequency of 7.4% from Masaka in 2014-2015 but not present in nearby Rukara. By 2018, 19.6% of isolates in Masaka and 22% of isolates in Rukara contained the mutation. Longitudinal monitoring is essential to inform control efforts. In Rukara, we sought to assess recent K13-561H prevalence changes, as well as for other key mutations. Prevalence of *hrp2/3* deletions was also assessed.

**Methods:** We genotyped samples collected in Rukara in 2021 for key artemisinin and partner drug resistance mutations using molecular inversion probe assays and for *hrp2/3* deletions using qPCR.

**Results:** Clinically validated K13 artemisinin partial resistance mutations continue to increase in prevalence with the overall level of artemisinin resistance mutant infections reaching 32% in Rwanda. The increase appears to be due to the rapid emergence of K13-675V (6.4%, 6/94 infections), previously not observed, rather than continued expansion of 561H (23.5% 20/85). Mutations to partner drugs and other antimalarials were variable, with high levels of multidrug resistance 1 (MDR1) N86 (95.5%) associated with lumefantrine resistance and dihydrofolate reductase (DHFR) 164L (24.7%) associated with antifolate resistance, but low levels of amodiaquine resistance polymorphisms with chloroquine resistance transporter (CRT) 76T: at 6.1% prevalence. No *hrp2* or *hrp3* gene deletions associated with diagnostic resistance were found.

**Conclusions:** Increasing prevalence of artemisinin partial resistance due to K13-561H and the rapid expansion of K13-675V is concerning for the longevity of artemisinin effectiveness in the region. False negative mRDT results do not appear to be an issue with no *hrp2 or hpr3* deletions detected. Continued molecular surveillance in this region and surrounding areas is needed to follow artemisinin resistance and provide early detection of partner drug resistance, which would likely compromise control and increase malaria morbidity and mortality in East Africa.

## Introduction

Malaria remains a global public health challenge. An estimated 247 million cases and 619,000 deaths occurred worldwide in 2021, with 95% of these cases and 96% of the deaths recorded from the WHO Africa region (1). The vast majority of deaths occur in children in Africa due to *Plasmodium falciparum* which accounts for 99% of malaria cases on the continent (2). The first-line treatment for *P. falciparum* infection is artemisinin-based combination therapies (ACTs), which combine a fast-acting artemisinin derivative with a longer-lasting partner drug that effectively eliminates any remaining parasites (3). But while ACT treatment is a cornerstone for test and treat strategies used throughout Africa, multiple artemisinin partial resistance (ART-R) mutations are now emerging that will likely reduce the effectiveness of treatment, hinder control efforts, and potentially further engender partner drug resistance and eventual ACT clinical failure (4–6).

ART-R manifests as delayed clearance after therapy and is most commonly mediated by various nonsynonymous propeller domain mutations in *pfkelch13* (*k13*) *gene* (*PF3D7_1343700*) (7). These mutations first emerged over a decade ago and have spread widely in Southeast Asia, along with subsequent partner drug resistance mutations. Together artemisinin and partner drug resistance causes clinical failure and recrudescence after ACT treatment (8). In the study initially characterizing ART-R in Africa, the Pfkelch13 561H mutation was found in 7.4% of samples from Masaka, Rwanda in 2015. Genomic analysis showed that 561H in Rwanda was genetically distinct in terms of flanking haplotypes from previously detected Southeast Asian 561H mutations, providing compelling evidence of a *de novo* local emergence in Africa (6). By 2018 in Masaka, the prevalence of 561H had increased dramatically to 19.6% and 561H was also found in 22% of samples from Rukara near the capital (9).

Further studies have shown that Eastern Africa has become the center of ART-R emergence with multiple validated K13 mutations, 469Y (Uganda), 561H (Rwanda), 622I (Eritrea and Ethiopia) and 675V (Uganda), independently emerging and spreading across borders to neighboring countries. In Uganda, clinical studies and detailed prevalence data over time show 675V and 469Y have increased over time and are now highly prevalent in multiple areas (4,10–14). In Eritrea and Ethiopia, 622I are increasing in prevalence (5,15). The 561H mutation has also appeared now in Uganda and Tanzania (16). In Rwanda, where 561H was first discovered, current data is more limited. The 561H mutation has been seen at appreciable prevalence in multiple sites (**Figure 1**, **Table 1**). In addition, in rare instances other candidate and validated mutations, such as a singular 675V isolate from the Huye district, are reported (9). However, concerted efforts at repeated or broader sampling are still lacking despite its initial discovery in Rwanda. Additional assessment is urgently needed to better understand emerging ART-R and the impact that 561H mutation may have on antimalarial therapy.

**Figure 1.**
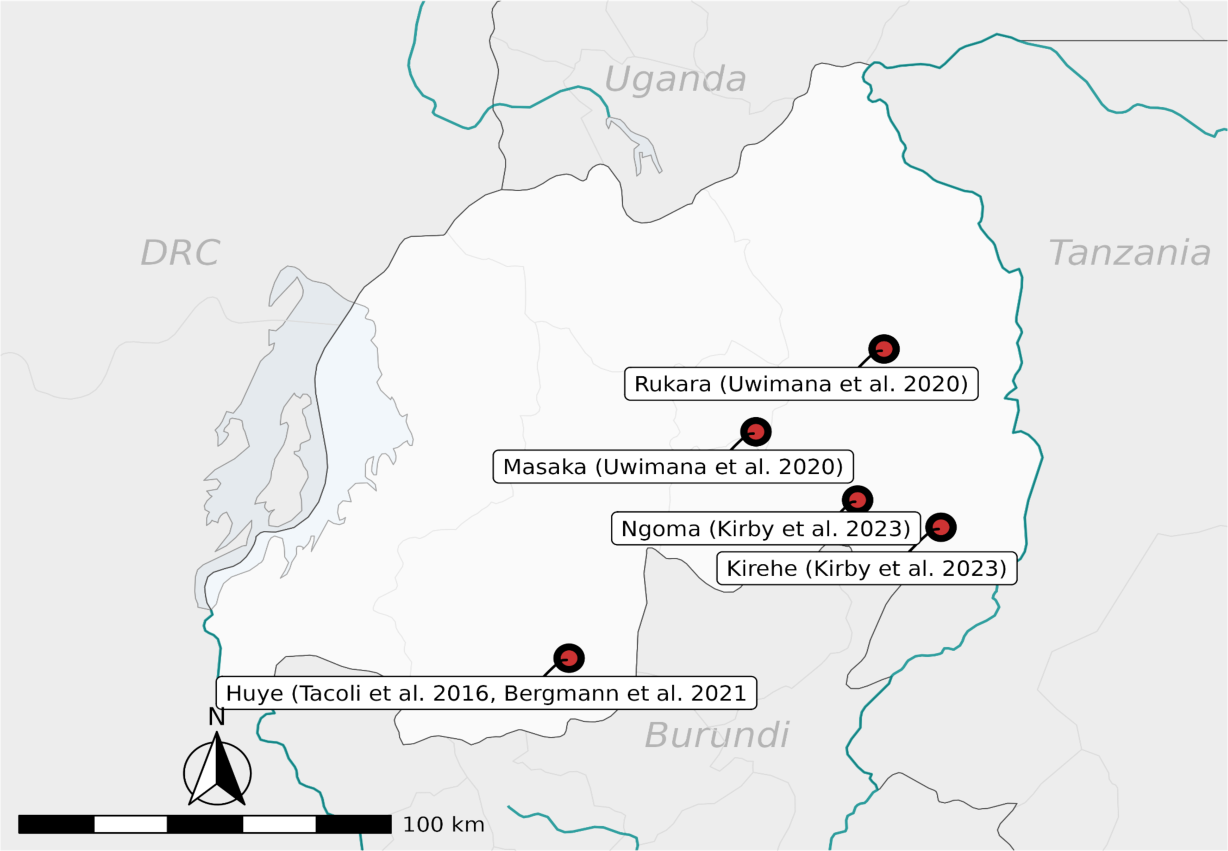
Distribution of Previous reports of K13 Mutations in Rwanda. The map illustrates the geographic distribution of research sites that have reported on K13 mutations, including Kirehe, Ngoma, Huye, Masaka, and Rukara.

**Table 1.** Prevalence of Polymorphisms Previously Reported in Rwanda.

| Mutation | Prevalence, location (year) | Citation |
| --- | --- | --- |
| 561H | 7.4%, Masaka (2015)<br>5.47%, Kirehe, (2014-2015)<br>2.47%, Ngoma (2014-2015)<br>19.6%, Masaka (2018)<br>22%, Rukara (2018)<br>4.5%, Huye District (2019) | (6,9)<br>(17)<br>(18) |
| 675V | Singular isolate, Huye District (2015)<br>4.5%, Huye District (2015) | (13)<br>(18) |

Another emerging challenge for test and treat strategies are *P. falciparum* histidine rich protein 2 and 3 (*hrp2/3*) gene deletions. *Hrp2* encodes the protein used in most malaria rapid diagnostic tests (RDTs) in Africa, thus deletion of the gene makes the parasite “invisible” or undetectable to RDTs. This diagnostic resistance has emerged and spread in the Horn of Africa and has the potential to emerge elsewhere. Little surveillance data for *hrp2/3* deletions exists in Rwanda. However, given areas of low transmission are of primary concern for the emergence and impact of these mutations, surveillance in Rwanda is likely important (19,20)

Here, we leveraged 273 samples, collected from uncomplicated malaria infections in Rukara, Rwanda during 2021 routine malaria programme clinical monitoring, to evaluate the status of molecular markers of antimalarial resistance 3 years from the last measures as well as to assess for the presence of *hrp2/3* gene deletions. Within our dataset, we found 3 of WHO’s 13 recognized validated K13 markers of resistance (R561H, P574L, and A675V) but none of their 9 candidate markers (21). Longitudinal monitoring of K13 mutations in Rukara helps to provide information about the changing patterns of partial artemisinin in resistance in Rwanda and provides valuable information for modelers and national control programs interested in studying the spread of polymorphisms that will impact test and treat strategies.

## Methods

### Patient samples

Dried blood spots (DBS) (n=273) were collected at Rukara Health Centre in 2021 with the intention of evaluating the performance of malaria diagnostics in Rukara, tracking the emergence of *hrp2/3* deletions, and monitoring drug resistance markers. Patients living in the catchment area of Rukara presenting clinical signs and symptoms of uncomplicated malaria with positive rapid diagnostic tests (RDTs) were recruited at the health center. RDT testing was done using the SD BIOLINE Malaria Ag P.f/Pan test to detect the histidine-rich protein II antigen of *P. falciparum* and pan *Plasmodium* lactate dehydrogenase of *Plasmodium* species in human whole blood. Recruited patients provided whole blood samples by intravenous draw that were used to confirm malaria infection through a malaria smear and the asexual parasite densities were estimated. This study was approved by the National IRB of Rwanda.

### Molecular Inversion Probe Genotyping

Blood spots were processed using the chelex method (22) to extract DNA and genotyped using molecular inversion probes (MIPs). We used the DR2 drug resistance MIP panel as previously detailed (23). This panel provides data from across *K13*, but also multiple other antimalarial resistance genes. Samples were demultiplexed using MIPTools software (available at https://github.com/bailey-lab/MIPTools) and variants were called using the freebayes setting with a minimum of 10 universal molecular identifiers per probe per sample. Further data cleaning and analysis was done in R version 4.1.2 using tidyverse and mapping using sf. Variants were called using the freebayes setting in MIPTools; all packages are detailed here: https://github.com/bailey-lab/Rwanda-DHS-2014-15. Confidence intervals for prevalence were determined using the Binomial exact calculation at https://sample-size.net/confidence-interval-proportion/.

### Molecular detection and determination of *hrp2/3* gene status

All samples were screened by a quantitative real time PCR assay for *Plasmodium falciparum* lactate dehydrogenase (*pfldh*) as previously described (24). A standard curve of mocked dried blood spot samples that used whole blood and cultured parasites (MRA-102, BEI resources, Manassas, VA) was generated. The mocked DBS was extracted in a similar fashion to the clinical samples, allowing us to estimate parasite density of infections after extraction based on real time PCR Ct value relative to the standard curve. For potential *hrp2/3* deletions, samples with a calculated parasitemia of 100 or more parasites per microliter were moved forward for *hrp2/3* deletion detection using a multiplexed real time PCR assay as previously described (25). The use of samples with higher parasitemia is necessary to reduce false positive *hrp2/3* deletion calls (26). Any sample that did not amplify at any gene, including *hrp2* or *hrp3*, was repeated before a genotype call was made.

## Results

We successfully genotyped 135 of the 273 samples. A summary of all antimalarial resistance polymorphisms in the samples is shown in **Table 2**. The 561H mutation was observed at 23.5% (95% CI: 15.0-34.0%) prevalence in 2021 compared to the 22% prevalence previously reported in Rukara in 2018 (9). Furthermore, the 675V mutation, which had previously not been reported on in the Rukara region, was found in 6.4% (95% CI: 2.4-13.4%) of isolates. No other validated or candidate ART-R polymorphisms were seen in the samples within the propeller domain, but three other non-synonymous polymorphisms were noted in the area (**Table 2**) (21). Furthermore, a common polymorphism K189T, which is outside of the propeller region, was found at 41.7%.

**Table 2:**
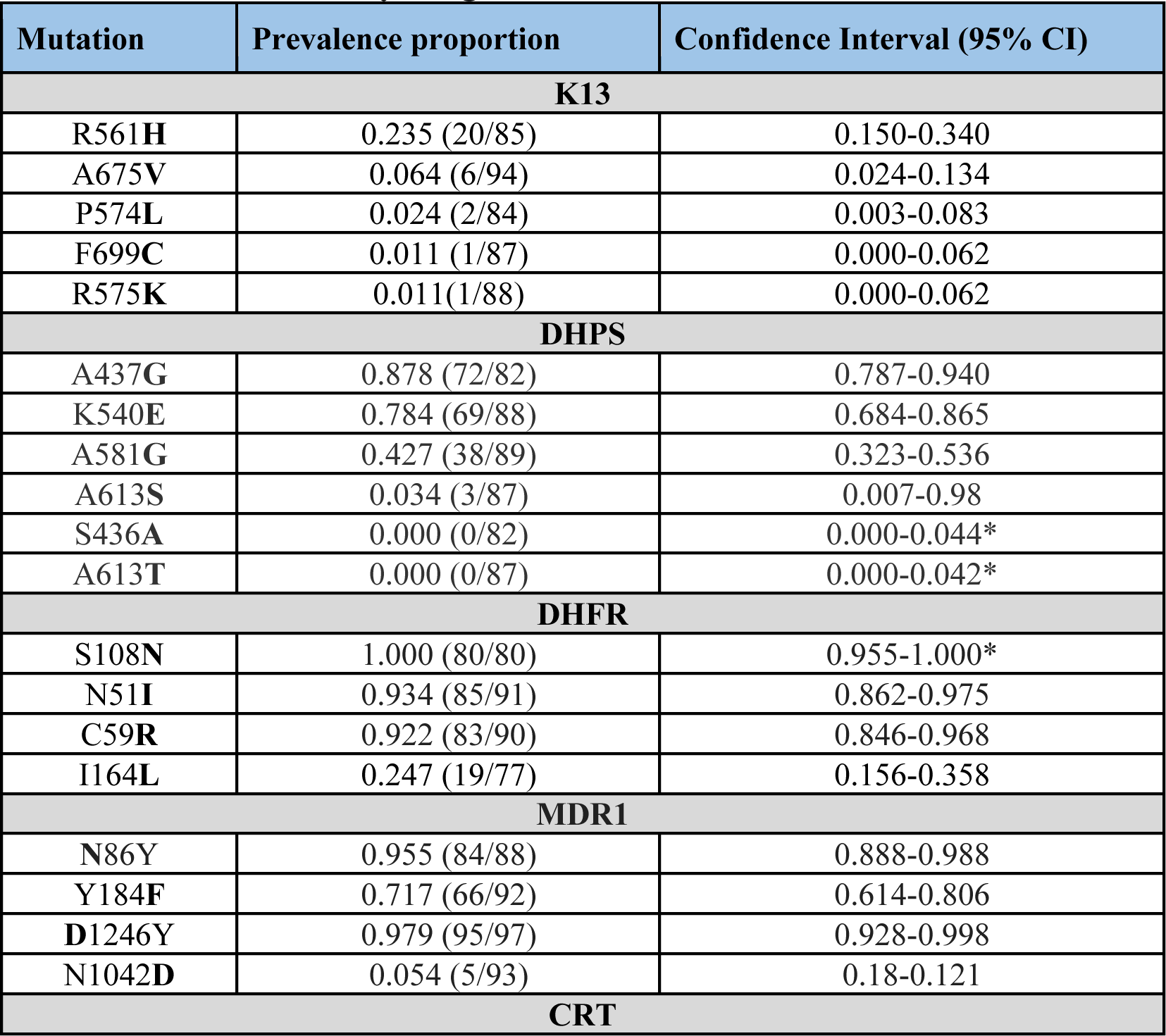
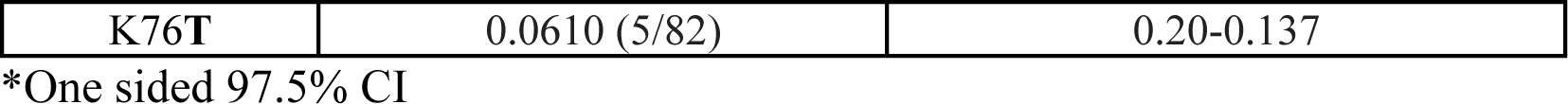
Prevalence of key drug resistance mutations.

In addition to K13, the MIP panel provides data on other antimalarial resistance polymorphisms. The primary ACT used in Rwanda is artemether-lumefantrine (AL). Having the wild type N86 amino acid of multidrug resistance protein 1 (MDR1) has been associated with tolerance to lumefantrine particularly in the context of the NFD (N86, 184F, D1246) haplotype. N86 was near fixation in the population with a prevalence of 95.5% (95% CI: 88.8-98.8%). The MDR1 NFD haplotype was seen in 72.3% (60/83) of isolates where all loci were available. The NFD haplotype did occur in samples with K13 mutations (Figure 2). Furthermore, resistance to chloroquine and amodiaquine have been linked to mutations in the chloroquine resistance transporter (CRT), particularly CRT 76T, which we found at a 6.1% prevalence among the samples. This mutation has been associated with better clinical response to artesunate-amodiaquine (ASAQ), which provides Rwanda with a backup drug if AL failure begins to happen (27,28).

**Figure 2.**
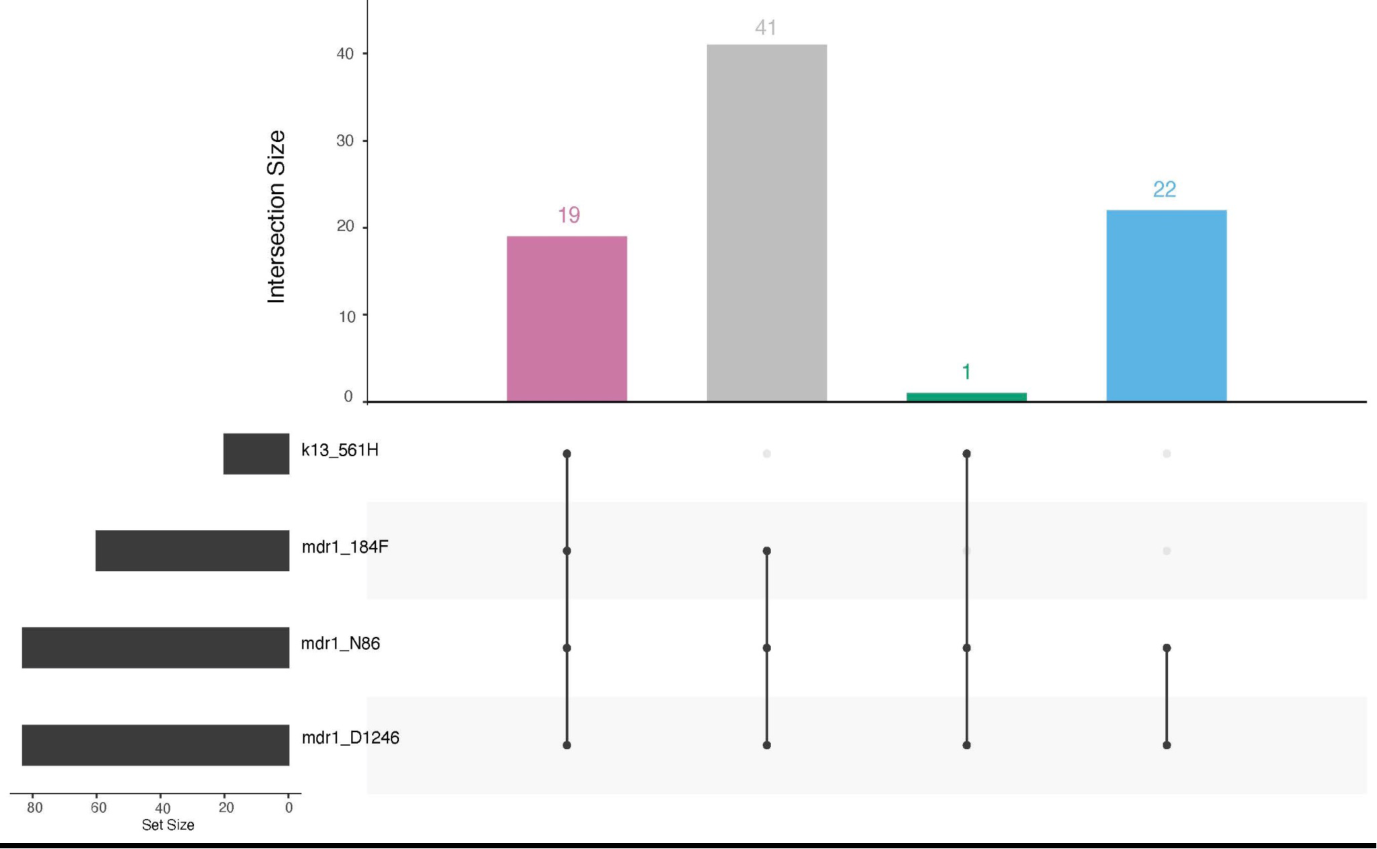
Upset plot of MDR1 and K13 mutations. A total of 82 samples had genotype calls at K13 R561H, MDR1 N86Y, MDR1 Y184F and MDR1 D1246Y. The majority of the parasites had the NFD haplotype in MDR1, with 31.7% (19/60) having the K13 561H mutation.

Resistance to SP occurs through sequential acquisition of mutations in two genes, dihydrofolate reductase (*dhfr*) and dihydropteroate synthase (*dhps*) (**Table 2**). Importantly, subsequent mutations in haplotypes of *dhfr*, such as 164L and *dhps*, such as 581G, are associated with high-grade pyrimethamine and sulfadoxine resistance, respectively, and significantly reduce the effectiveness of SP used for chemoprevention. Here we see the DHFR 164L mutation at a prevalence of 24.7% (95% CI: 15.6-35.8%) and the DHPS 581G mutation at a prevalence of 42.7% (95% CI: 32.3-53.6%) (**Table 2**). The DHPS 581G mutation occurred in 45.8% (38/83) of samples where data was available for DHPS 581G, 437G, and 540E mutations (Figure 3).

**Figure 3.**
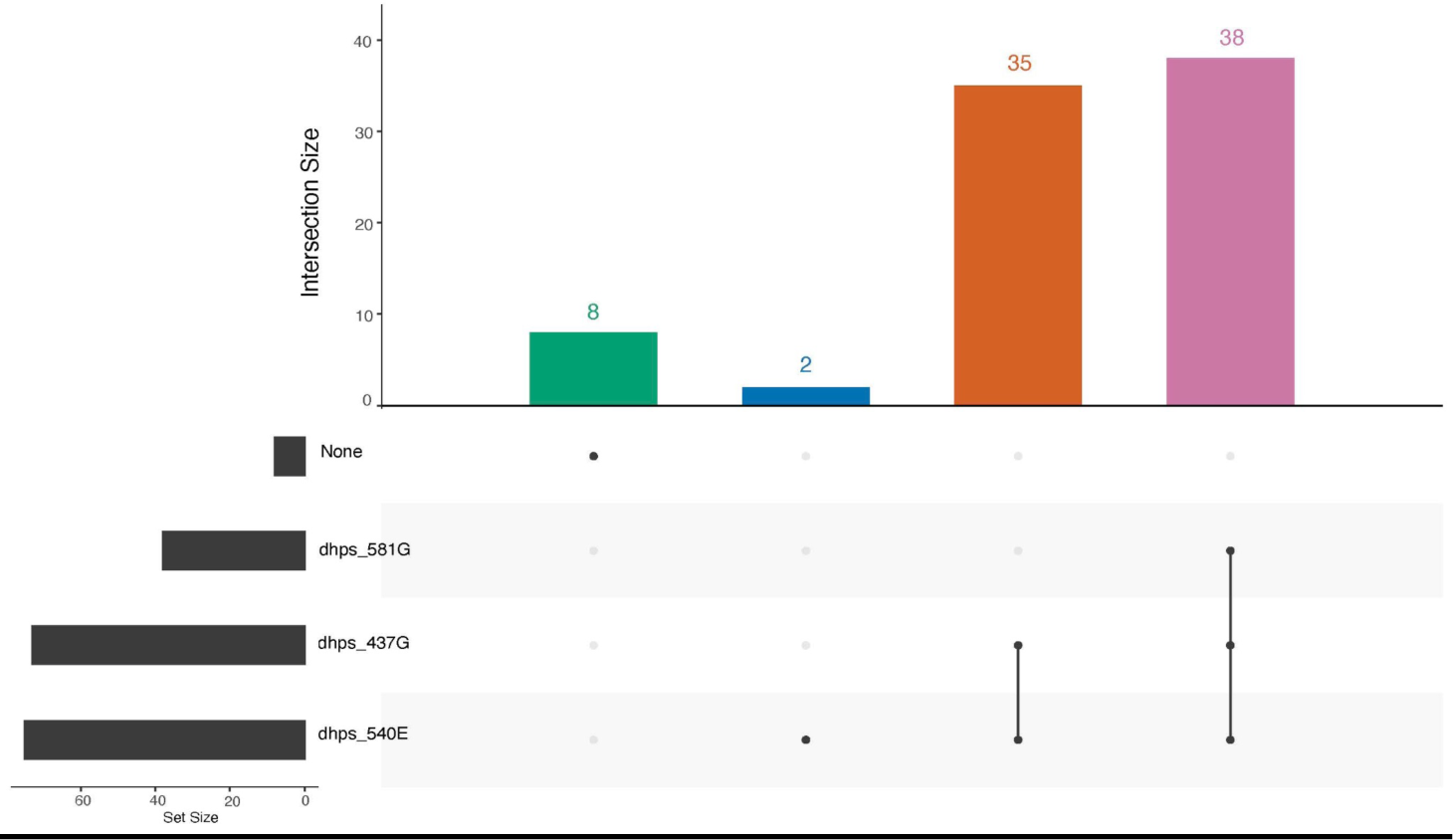
Upset plot of DHPS mutations. A total of 83 samples had genotype calls at DHPS A437G, K540E, and A581G and 45.8% (38/83) of the infections had all three mutations, while 88.0% (73/83) had both DHPS 437G and 540E.

To assess diagnostic performance and evaluate for the presence of *hrp2/3* deletions, we examined 274 samples through RDT, microscopy, and qPCR to assess their positivity; it was observed that 8 samples (Figure 4) were RDT negative, despite exhibiting positive microscopy and positive qPCR results indicating potential *hrp2/3* deletions in the sample. Of these eight, four had high enough parasitemia to assess for deletions by real time PCR, of which none contained deletions. Using qPCR as the gold standard (**Table 3**), the RDT had an NPV of 0.71 and a PPV of 0.97, while the blood smear microscopy hadd an NPV of 0.70 and a PPV of 0.88.

**Figure 4.**
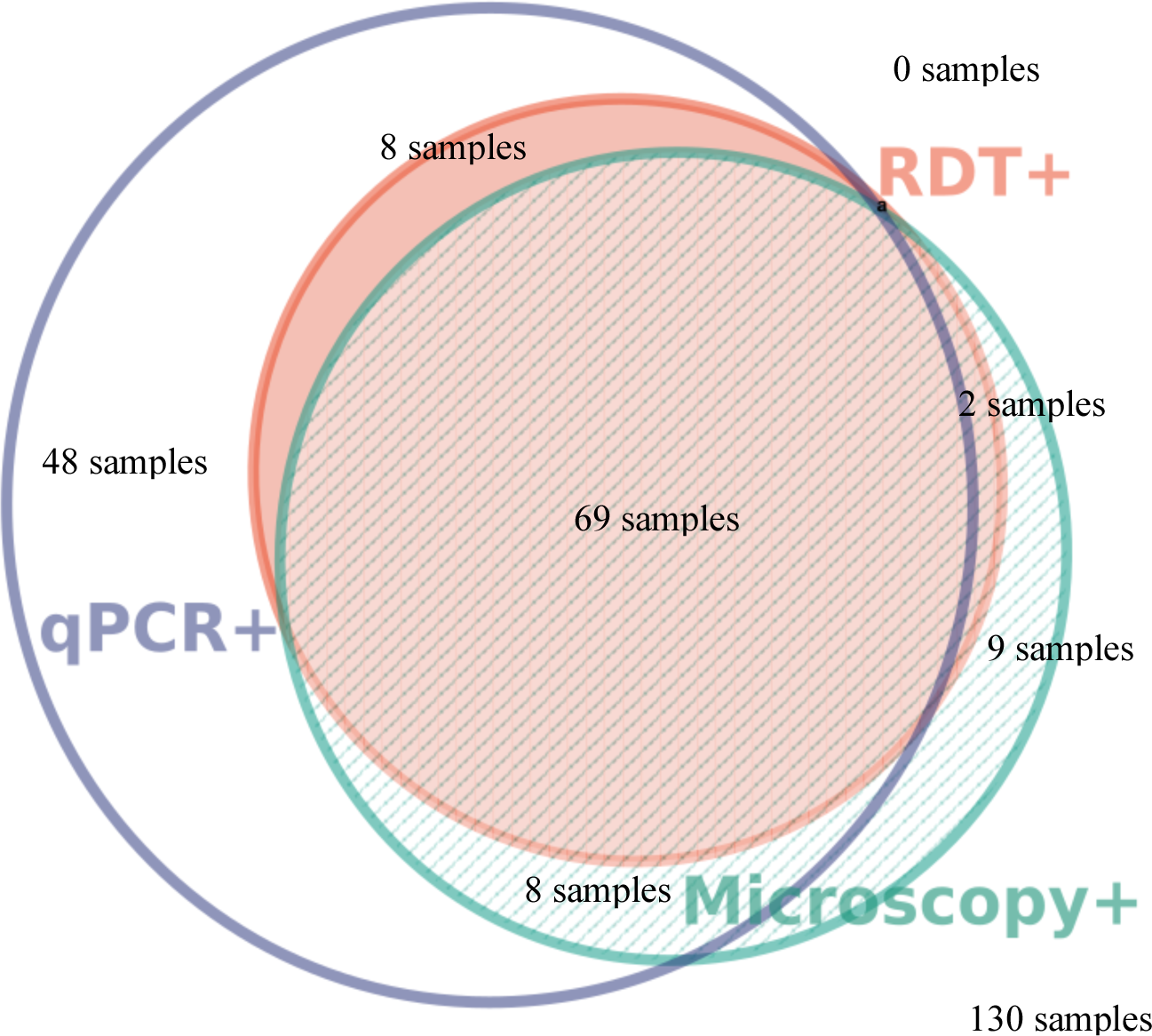
Proportional Euler Venn Diagram Comparing the Three Utilized Diagnostic Methods. 130 of the 274 samples were negative for all three tests and are excluded from the venn diagram (29)

**Table 3:**
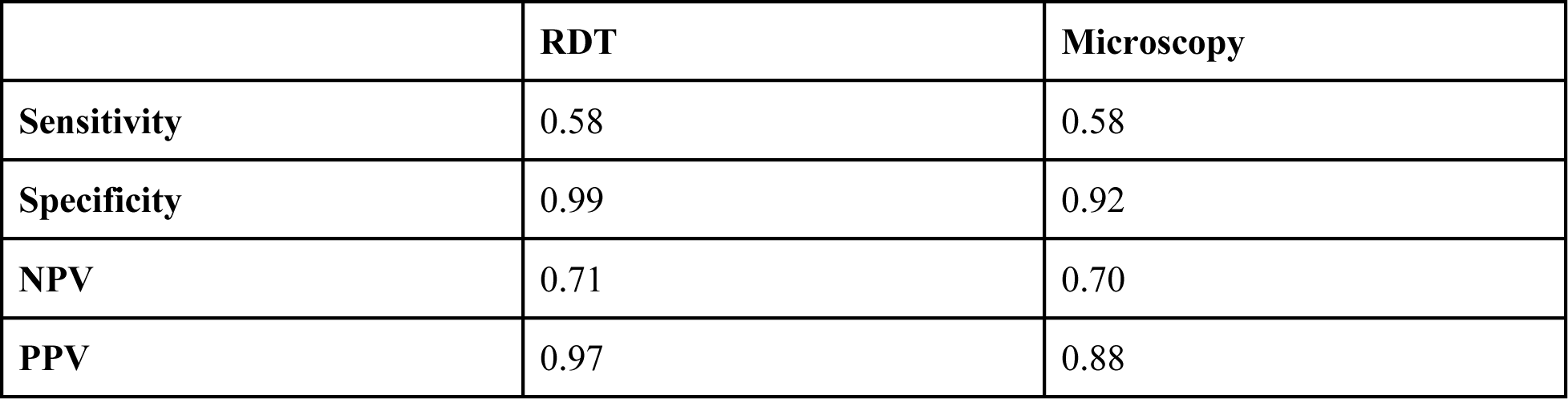
Statistics on RDT and Microscopy using qPCR as a gold standard.

|  | RDT | Microscopy |
| --- | --- | --- |
| **Sensitivity** | 0.58 | 0.58 |
| **Specificity** | 0.99 | 0.92 |
| **NPV** | 0.71 | 0.70 |
| **PPV** | 0.97 | 0.88 |

## Discussion

Our study provides additional insight into the ongoing emergence of artemisinin partial resistance by providing longitudinal data from one of the sites in Rwanda with early descriptions of K13 mutations. Importantly, we report the 675V mutation, previously found in Uganda (Conrad et al., 2023). Meanwhile, as previously said, 561H appears to be relatively stable in the population (23.5% in 2021 compared to 22% from 2018 (9), and falls within the range modeled for the 561H allele frequency for Rwanda in 2021 (30). However, the overall level of all validated K13 mutants has increased to 32% with the arrival of the 675V mutation. The 675V mutation has reached high levels in neighboring Uganda and the emergence in Rukara likely represents spread across the border given the high-levels of spread within Uganda itself (4,10). However, an independent origin can not be ruled out without additional genomic analysis of examining the haplotypes flanking the *k13* gene. In the end, the overall increase in ART-R among parasites is concerning for malaria control in the region.

Beyond K13, we find evidence for mutations important for understanding partner drug effectiveness. The combination of high levels of MDR1 N86 and low CRT 76T are reassuring that Rwanda has an effective backup ACT with ASAQ if their first line should fail. AL continues to have good clinical effectiveness, but high prevalence of the MDR1 N86 mutation is concerning. The MDR1 NFD haplotype has been linked to reduced effectiveness towards lumefantrine (31). The current study found N86 in 95.5% (95% CI: 88.8-98.8%), 184F in 71.7% (95% CI: 61.4-80.6%) and D1246 in 97.9% (95% CI: 92.8-99.8%) of isolates. The NFD haplotype was found in 72.3% (60/83) of samples where all loci were available and this haplotype did occur with K13 mutations (Figure 2). This aligns with the results of a recent study in Ethiopia, where the NFD haplotype was found in 83% of the 609 samples collected and co-occurred with the K13 622I mutation 98% of the time (5)).

SP is routinely used as an intermittent preventive treatment in pregnancy (IPTp) for malaria as well as for seasonal malaria chemoprophylaxis (SMC) and intermittent preventive treatment during infancy (IPTi) in Africa. Historically drug pressure from SP drove the sequential mutations in DHFR and DHPS, but continued pressure from some SP use and activity against malaria parasites by co-trimoxazole (trimethoprim-sulfamethoxazole) may have impacted their prevalence (32). Currently, Rwanda is not using SP for IPTi as resistance markers are found above WHO thresholds for continued use (21). Escalating prevalence of SP resistance markers endangers the efficacy of malaria prevention during pregnancy regionally given evidence of spread (33). In particular, the DHPS 581G mutation and DHFR 164L mutations are associated with high level resistance to SP (34). The latter, DHFR 164L, known to confer heightened resistance to antifolates, had previously been seen with a prevalence of 11% in Rukara between 2001 and 2006 (35). Its prevalence was much higher in areas of Uganda, where a 35% prevalence was reported in Fort Portal in 2013 (36). The impact on clinical efficacy of chemoprevention and the evidence of regional spread highlight the need for continued monitoring of these mutations even in countries like Rwanda where IPTp is not used routinely.

DHFR and DHPS mutations were common in the study area (**Table 2**). Notably, we identified DHPS 540E at a prevalence of 78.4% (95% CI: 68.4-86.5%), above the threshold set by the WHO (>50%) to halt SP-IPTi (37). The DHPS 581G mutation occurred in 42.7% (95% CI: 32.3-53.6%) of isolates, with 45.8% (38/83) of samples, where data from all polymorphisms is available, showing the 437G, 540E, and 581G mutation together (Figure 3). The presence of late high level antifolate resistance mutations in the region are very concerning. DHFR 164L was found at a prevalence of 24.7% (95% CI: 15.6-35.8%). Regional spread of these mutations is a concern and high levels of these mutations have recently been described in North-West Tanzania (16). An interesting finding is the presence of the DHPS 613S mutation found at a prevalence of 3.4% (95% CI: 0.7-9.8%); this is a mutation typically found in West Africa (38). However, they have been reported at low frequency as close as neighboring Democratic Republic of the Congo (39). Overall, this data aligns with previously reported trends in DHFR and DHPS mutations in Rukara where 75% of the isolates tested had three mutations in DHFR and two or three in the DHPS gene (35).

The emergence of K13 675V in Rukara is concerning, as it shows the spread of the mutation from Uganda, or potentially an independent origin. While the mutation was reported in the Huye District of southern Rwanda in 2015, no studies have detected it at an appreciable frequency in Rwanda. More concerningly, 675V appears to have increased in frequency more rapidly than 561H suggesting the possible fitness advantage compared to 561H. This advantage may be attributable enhanced survival following treatment with ACT and/or better survival and growth in the absence of drugs. While there are also other K13 mutations that we found in our dataset that haven’t been reported on in Rukara before, namely P574L, F699C, and R575K, their prevalences are all less than 0.03 (3%) and unvalidated which raises questions about their significance and calls for further investigation to determine their impact and potential implications in Rukara.

While we have been able to provide new data on the longitudinal changes in K13 mutation in Rukara in this study, we are limited in many aspects. These samples represent a convenience sample of participants enrolled in a study not meant to monitor antimalarial resistance and thus may not be population representative. The geographic scale is also small, and though any longitudinal data for molecular surveillance is valuable, broadly geographically representative sampling reflects the best means for understanding antimalarial resistance trends (4,10,33).

*Hrp2/3* deletions were not detected in samples discordant by RDT compared to PCR and microscopy. This is reassuring that RDTs that were negative were not due to these deletions, but can be due to other reasons such as lot-to-lot variation, operator error, and poor storage but in this case it is likely due to the fact that HRP2 is not a perfect correlate with parasitemia--i.e. every parasitemia >100 p/uL may not have enough detectable HRP2 (20,40).

The results of this study support that artemisinin partial resistance, partner drug resistance, and resistance to antimalarials used for antifolate chemoprophylaxis are all emerging and will pose imminent challenges for malaria control programs in Africa. Longitudinal molecular surveillance is needed for addressing these concerns in a timely manner and to help inform policy. Effective molecular surveillance can help rapidly inform where either *in vitro/ex vivo* assessment resistance or therapeutic efficacy studies can be targeted. Emergence of *hrp2/3* deletions in Africa may also eventually pose a challenge outside of the Horn of Africa.

Fortunately *hrp2/3* deletions do not appear to be a problem in Rwanda at this time. Together, this information provides valuable data for the national malaria control program and for modelers and other scientists interested in studying the emergence and spread of antimalarial resistance.

## Data Availability

All data produced in the present study are available upon request to the authors.

## Acknowledgments

We thank the participants of the study. The following reagent was obtained through BEI Resources, NIAID, NIH: *Plasmodium falciparum*, Strain 3D7, MRA-102, contributed by Daniel J. Carucci.

